# Circulating proteins altered in response to the Dietary Approaches to Stop Hypertension (DASH) diet suggest underlying molecular mechanisms and long-term health benefits

**DOI:** 10.64898/2026.07.22.26358749

**Authors:** Hyunju Kim, Jennifer A. Brody, Guanghao Qi, Ting Ye, Rizwan Kalani, Lawrence J. Appel, Casey M. Rebholz, Neil M. Davies, James S. Floyd

**Author notes:** **Address Correspondence to:** Hyunju Kim Department of Epidemiology University of Washington 3980 15th Ave NE Seattle, WA 98105.

## Abstract

The Dietary Approaches to Stop Hypertension (DASH) diet reduces blood pressure and cholesterol. However, the mechanisms underlying these effects are unclear, and no randomized studies have evaluated the long-term benefits on health outcomes such as coronary artery disease (CAD) or type 2 diabetes (T2D). We performed a series of Mendelian randomization analyses of 71 serum proteins perturbed by the DASH diet in previous randomized controlled feeding studies to understand their potential mechanistic role on health outcomes. Four proteins (ANGPTL3, INHBC, PCOLCE, PLXNB2) had causal evidence of beneficial effects on risk factors that aligned with diet-induced changes in protein levels. Missense variants in INHBC and PLXNB were associated with lower risks of CAD and T2D respectively, and these effects are directionally concordant with DASH diet-induced changes in protein levels, providing causal evidence that these proteins influence disease risk. Combining molecular phenotyping in randomized interventional studies with human genetics evidence identified molecular regulators of the long-term cardioprotective effects of the DASH diet.

## Main

Randomized, controlled, feeding studies have provided evidence of the casual effects of specific dietary patterns on health, shaping national dietary guidelines.^1^ However, such trials are often short-term (several weeks) and focus on biomarkers and intermediate health outcomes such as blood pressure, due to the high cost and the impractical sample sizes needed to detect effects on long-term cardiovascular outcomes, such as coronary artery disease (CAD) and type 2 diabetes (T2D). As a result, high-quality evidence about the potential long-term health benefits (or harms) of dietary interventions and underlying causal biological mechanisms has remained difficult to obtain. We recently described an approach that leverages results from large-scale human genetic studies of plasma proteomics and various health outcomes to support inference about long-term health benefits and their molecular mediators, which we now apply to a dietary intervention.^2^

Plasma proteins transport nutrients and mediate metabolic reactions, immune responses, and cellular responses,^3^ functions relevant for digestion, absorption, and metabolism of food. Unlike dietary intake, proteins have strong genetic determinants, and most proteins detected by widely-used affinity-based proteomics platforms have protein quantitative trait loci (pQTL) near the genes encoding the proteins (*cis*-pQTLs).^4^ We hypothesized that proteins affected by diets and supported by causal genetic evidence may mediate beneficial health effects, facilitating discovery of causal mechanisms and helping to establish their long-term health effects that contemporary feeding studies are not powered to evaluate.

The Dietary Approaches to Stop Hypertension (DASH) diet is high in fruits, vegetables, and low-fat dairy; includes a variety of protein sources; and limits red and processed meats and sugar-sweetened beverages.^5^ The DASH randomized, controlled feeding trial found that individuals who were provided with the DASH diet for 8 weeks had a reduction in systolic blood pressure (SBP) by 5.5 mm Hg, diastolic blood pressure (DBP) by 3.0 mm Hg,^6^ low-density lipoprotein (LDL)-cholesterol (C) by 0.28 mmol/L, and total cholesterol (TC) by 0.35 mmol/L^7^ compared to those who were provided with the control diet (typical American diet). Similar results were reported from the DASH-Sodium randomized, controlled feeding trial.^8^ We recently found that the DASH compared to the control dietary patterns tested in the DASH and DASH-Sodium feeding trials affected serum levels of 71 proteins, after correcting for multiple comparisons (false discovery rate <0.05) (**Figure 1A**).^9^ We evaluated this set of candidate proteins in a series of reverse and forward Mendelian randomization analyses,^4^ which uses genetic variation and the random allocation of alleles to estimate causal effects of one trait on another.^10^ Specifically, we evaluated whether genetically determined levels of these DASH diet-related proteins have causal effects on cardiovascular risk factors [SBP, DBP, LDL-C, and TC], and clinical endpoints [CAD, T2D, ischemic stroke, and heart failure].

**Figure 1.**
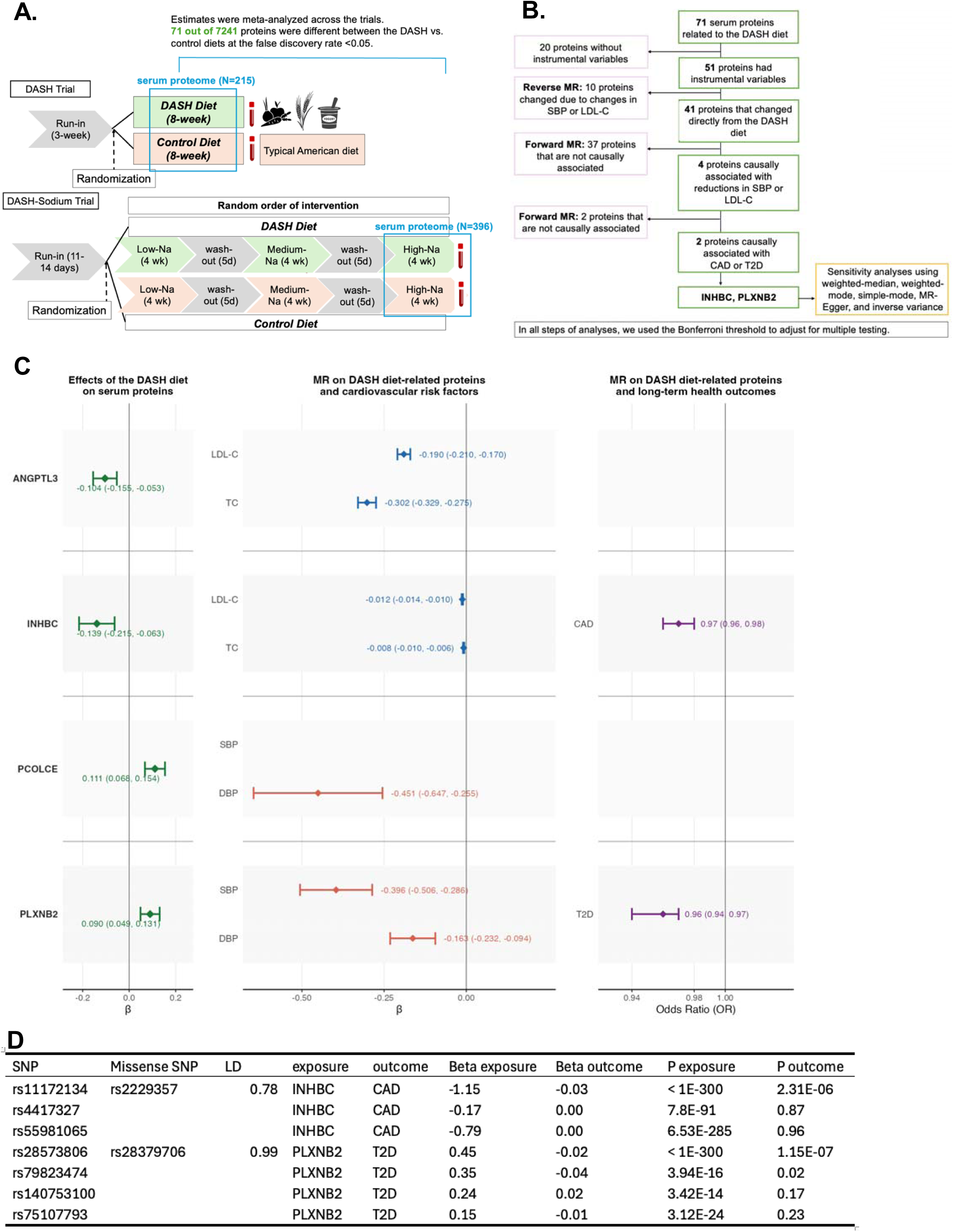
(A) Design of the DASH and DASH-Sodium proteomics feeding studies, (B) flowchart of analysis, and (C) Mendelian Randomization analysis estimating the causal effects of DASH diet-related proteins, cardiovascular risk factors, and long-term health outcomes. All estimates are expressed as per standard deviation (SD) change in protein level in the direction of the observed protein effect. For proteins with positive observed effects, β and odds ratios represent the outcome change per SD increase in protein levels; for proteins with negative observed effects, β and odds ratios represent the outcome change per SD decrease in protein level. **(D)** Mendelian randomization estimates of the causal effects of INHBC and PLXNB2 on CAD and T2D, respectively. For each protein, we searched the associated instrumental variant SNPs for missense variants in the corresponding gene; this identified rs2229357 as a missense variant in INHBC, and rs28379706 as a missense variant in PLXNB2. No candidate proteins were causally associated with incident heart failure or ischemic stroke. CAD, coronary artery disease; DASH, Dietary Approaches to Stop Hypertension; EAF, effect allele frequency; LDL-C, low-density lipoprotein-cholesterol; T2D, type 2 diabetes

Of the 71 candidate proteins, which were assayed with SomaScan v4.1, 51 proteins had *cis-*pQTLs from previous studies that used older versions of the SomaScan proteomics platform (**Figure 1B**). Because this work is attempting to identify molecular changes that are a direct result of dietary intake, we excluded an additional 10 proteins whose perturbations may be explained by causal effects of SBP, DBP, LDL-C, and TC on the proteins themselves (p <0.05/51 proteins = 9.80 × 10^-4^) (**Supplemental Table 1**).

For the remaining 41 proteins, we genetically-instrumented the directional changes in proteins levels caused by the DASH diet and then we evaluated these causal effects on blood pressure and cholesterol phenotypes, which were directly evaluated in the clinical trials, and long-term cardiovascular health outcomes, which were not. After correcting for multiple comparisons (p <0.05/41 = 1.22 x 10^-3^), we identified 4 proteins [angiopoietin-related protein 3 (ANGPTL3), inhibin beta C chain (INHBC), procollagen C-endopeptidase enhancer 1 (PCOLCE), and plexin-B2 (PLXNB2)] whose genetically instrumented effects on blood pressure and cholesterol aligned with the results of the DASH trials (**Supplemental Table 2; Figure 1C**). Genetically-determined lower levels of ANGPTL3 and lower INHBC were causally associated with reductions in LDL-C and TC (range of β=-0.003 to -0.302), whereas higher levels of PCOLCE and higher PLXNB2 were causally associated with reductions in SBP and DBP (PCOLCE associated with DBP only) (range of β=-0.163 to -0.396). These results were consistent in sensitivity analyses which used weighted-median, weighted-mode, simple-mode, MR-Egger, and inverse variance (**Supplemental Table 3**).

Two of these proteins, INHBC and PLXNB2, had evidence of causal effects on long-term clinical outcomes that were driven by instrumental variables in linkage disequilibrium with missense variants (rs2229357 and rs28379706). (**Figure 1D**). The rs2229357 A allele in *INHBC* resulted in lower SomaScan-measured protein levels and associated with lower LDL-C, TC, and CAD risk (OR 0.97, 95% CI: 0.96–0.98); the rs28379706 C allele in *PLXNB2* resulted in higher SomaScan-measured proteins levels and associated with lower SBP, DBP, and T2D risk (OR 0.96, 95% CI: 0.94–0.97). To determine whether these missense variants reflect true changes in protein abundance or affinity-based binding effects, we examined peptide-level associations in an independent mass spectrometry dataset (SEER Proteograph, n=200, Cardiovascular Health Study). For INHBC, the SNP rs11172134 (r^2^=0.78 with the missense variant rs2229357) was strongly associated (p=6.02 × 10^-18^) with only the peptide altered by the missense change (ANTAAGTTGGGSCCVPTA[R->Q]), while no other peptides were significantly associated with the SNP; excluding this peptide from the aggregate protein-level signal, computed across all peptides, weakened the association from p=1.05 × 10^-5^ to p=0.0455. For PLXNB2, the SNP rs28573806 (r^2^=0.99 with the missense variant rs28379706) was not associated with overall protein abundance, but the single peptide altered by the missense change (SSGGPGAGLCLFPLD[K->E]) was strongly associated with the SNP (p=1.7 × 10^-7^), while no other peptides were significant associated. Accordingly, these MR associations likely reflect the causal relevance of altered protein structure to disease, rather than the effect of diet-induced changes in protein levels. No candidate proteins were causally associated with incident heart failure or ischemic stroke outcomes.

Integrating blood proteomics and Mendelian randomization, we identified molecular mechanisms linking a specific healthful dietary pattern (DASH diet) to beneficial effects on cardiovascular risk factors and outcomes. One well-established lipid-lowering drug target was identified by this approach (ANGPTL3), and as well as two novel proteins that may capture some of the long-term health effects of a dietary intervention. For both INHBC and PLXNB2, the MR instruments were missense variants, meaning the causal estimates reflect altered protein structure and function rather than circulating abundance alone. How the causal effects of protein isoforms relate to the effects of concentrations is unknown for these proteins. Nonetheless, the directional concordance between the diet-induced changes in protein levels observed in the feeding studies and the clinical associations of these missense variants supports the hypothesis that INHBC and PLXNB2 lie on the causal pathway linking the DASH diet to long-term cardiovascular and metabolic outcomes.

ANGPTL3, produced in hepatocytes, is recognized as a therapeutic target to lower LDL-C and triglycerides.^11^ In a predominantly European ancestry cohort, rare naturally occurring loss-of-function variants in *ANGPTL3* was associated with significantly lower levels of triglycerides, LDL-C, and reduced risk of coronary heart disease.^12^ ANPTL3 inhibition is thought to reduce LDL-C levels by 1) activating lipoprotein lipase, increasing hydrolysis of VLDL cholesterol, and improving plasma triglyceride clearance, or 2) elevating endothelial lipase, thereby driving compositional remodeling of VLDL, and facilitating removal of VLDL remnants in the absence of LDLR.^13^ The DASH diet reduced serum ANGPTL3 levels, highlighting a potential molecular mediator through which the DASH diet reduces LDL-C levels.

INHBC, expressed predominantly in the liver, is part of the transforming growth factor (TGF)-superfamily which plays a crucial role in regulating metabolic homeostasis^14^ and cholesterol metabolism.^15^ Although the effects of INHBC on cardiometabolic traits have not been explored extensively, this protein is a ligand of the TGF-receptor activin receptor-like kinase 7 (ALK7) which is highly expressed in adipose tissue.^16^ A recent study using bidirectional Mendelian randomization and immortalized human abdominal and gluteal adiposity found evidence of cross-talk between liver-derived INHBC and ALK7 in white adipose tissue.^17^ In human abdominal and gluteal adipocytes, INHBC binding to ALK7 triggered SMAD2/3 phosphorylation and reduced adrenaline-stimulated lipolysis.^17^ In addition to this local effect, INHBC was suggested to affect hepatic lipid metabolism through broader endocrine actions on white adipose tissue by decreasing lipid turnover.^17^ Adipose tissue lipolysis may be a key pathway through which the DASH diet reduces LDL-C and incident CAD. Another mechanism by which the DASH diet can lower LDL-C and total cholesterol is through reduced intake of atherogenic cholesterol such as saturated fat. Clinical trials have reported consistently that greater intake of saturated fat, especially if saturated fat replaces polyunsaturated fat, leads to increases in LDL-C.^18^

Similar to INHBC, a missense variant in PLXNB2 was associated with reduced blood pressure and reduced risk of T2D. PLXNB2 which serves as receptors for semaphorin 4D, is primarily known to direct neural cell growth during brain development, and secondarily play a role in developing other organs, such as kidney.^19^ PLXNB2 is expressed in a wide range of nonneuronal adults cells, including in endothelial cells, pancreatic islets, and medulla and cortex of the kidneys.^20^ Prior Mendelian randomization analysis found that greater genetically determined PLXNB2 levels were inversely associated SBP or DBP, and a lower risk of incident T2D.^21^ The potential role of PLXNB2 in cardiovascular disease has not yet been explored. Mechanistic studies are warranted to clarify the regulatory role of circulating PLXNB2 in influencing blood pressure and T2D risk.

Genetically determined higher levels of PCOLCE was associated with reductions in diastolic blood pressure. PCOLCE, expressed abundantly in collagen-rich tissues, such as skin and lower levels in other tissues (in the heart, kidney, and skeletal muscle) accelerates the conversion of procollagen to mature collagen in extracellular matrix by increasing the activity of bone morphogenetic protein-1/tolloid-like proteinases (BTPs), an enzyme that breaks down C-terminal propeptides.^22^ PCOLCE has been extensively studied in relation to fibrotic disorders, including cardiac fibrosis.^23^ Recently, animal models found that reactive oxygen species/DNA damage/c-Fos/C-Jun pathway leads to greater PCOLCE production in brown adipose tissue, which can further increase cardiac fibrosis and diastolic dysfunction.^24^ The DASH diet, high in phytosterol and polyphenols, can manage reactive oxygen species by improving oxidative stress and increasing the release of vasodilator such as prostacyclin.^25^ In fact, a systematic review of randomized controlled trials found that the DASH dietary pattern lowered lipid peroxidation, and increased glutathione.^26^

This study has several strengths. We used the power of randomization in two feeding studies to identify serum proteins that are impacted by the DASH diet, a widely recommended dietary intervention. A different kind of randomization – from the random assortment of alleles at gametogenesis – and large external datasets were used to study the causal effects of these proteins on short- and long-term cardiovascular outcomes. The DASH and DASH-Sodium feeding studies are an ideal setting to discover proteins responsive to dietary intake, given that participants were provided all cooked meals, including snacks, and had excellent adherence to the study diets as documented with by serum and urine biomarkers.^6,8^ Using this approach, from a pool of candidate biomarkers of the DASH dietary pattern, we distinguished markers that causally contribute to cardiovascular health thus merit further mechanistic study, and evaluated the effects of the DASH diet on hard clinical endpoints.

Several limitations are worth noting. First, we could not evaluate all 71 proteins responsive to the DASH dietary pattern, because our analyses were restricted to the proteins with strong pQTLs. In the DASH and DASH-Sodium feeding studies, we used the more expansive SomaScan v4.1, whereas the previous GWAS used SomaScan v4. Strong pQTLs could be identified as future GWAS start to incorporate updated proteomic assays. Second, evaluating the violations of the assumption of MR, horizontal pleiotropy is challenging. However, broadly consistent results across multiple complementary MR methods in sensitivity analyses speak to the robustness of our findings. Third, pQTLs and outcome data sets came from largely European ancestry populations, which limits the genetic diversity of these analyses. Nonetheless, more than half of the study participants in the DASH and DASH-Sodium feeding studies were minority populations.

In summary, we establish and illustrate a framework to identify causal proteins amenable to dietary change that contribute to cardiovascular benefits, and triangulate potential long-term effects of these causal proteins on CAD and T2D. This approach can be applied to future feeding studies to elucidate the underlying mechanisms of dietary intake and health outcomes, and predict whether short-term dietary changes translate into long-term health benefits.

## Methods

A series of reverse and forward Mendelian randomization analysis evaluated causality between 71 DASH diet-related proteins, assessed with aptamers (SomaLogic, Boulder, Colorado), and cardiovascular risk factors, and clinical end points (flowchart of analysis presented in **Figure 1B**) using debiased inverse-variance weighted approach.^10,27^ First, we conducted reverse two-sample Mendelian randomization to exclude proteins whose levels may have changed as a consequence of changes in cardiovascular risk factors rather than directly in response to the diet. In these analyses, the exposures were SBP, DBP, LDL-C, and TC, and the outcome was the level of each protein. Genetic instruments for each cardiovascular risk factor were identified from the largest available genome-wide association studies of European ancestry.^28,29^ For proteins, we used pQTLs obtained from deCODE at the genome-wide significance level (p < 5.0 × 10^-8^ ). A set of IVs was identified by clumping at a threshold of r^2^ <0.001 and 10 KB apart using the European linkage disequilibrium (LD) reference panel from the 1000 Genomes Project to obtain independent variants.^30^ To ensure that the instruments reflected variation in the cardiovascular risk factor rather than in the protein itself, we excluded any IVs that were *cis* to the protein being evaluated (within 1 MB of the gene encoding the protein). Additionally, we removed proteins that were directionally discordant with the main effects of the DASH diet on cardiovascular risk factors (i.e., proteins causally associated with an increase in SBP, DBP, LDL-C, or TC).

Second, we conducted three-sample forward Mendelian randomization on the remaining proteins. We selected genetic variants associated with each protein at genome-wide significant levels (p < 5.0 × 10^-8^) using *cis*-pQTLs (1 MB +/- transcription start site) reported in a large European ancestry study,^4^ and clumped to independent variants (r^2^<0.001) using the same criteria as described above. To reduce the possibility of winner’s curse and to increase precision, we then identified the associations of each SNP with each protein using summary data from deCODE. These instruments were then used to evaluate causal effects on SBP, DBP, LDL-C, and TC.^28,29^ We additionally attempted to falsify the exclusion restriction assumption by removing variants with stronger associations with other proteins in the ARIC pQTL Study, ensuring that the instruments were specific to the protein of interest. Lastly, for only those proteins that were causally associated with any of SBP, DBP, LDL-C, and TC, proteins which had ‘anchoring’ evidence on short-term improvement in cardiovascular risk factors, we used Three Sample MR to test their causal association with clinical end points (CAD, T2D, stroke, and heart failure). Potential IVs for proteins were identical to previous analysis. The summary data for outcomes were obtained from large European ancestry meta-analyses GWAS of CAD,^31^ T2D,^32^, stroke,^33^ and heart failure.^34^ In all steps, we applied the Bonferroni threshold to correct for multiple testing. For the proteins that were causally associated with clinical endpoints, we conducted multiple sensitivity analyses which are robust to invalid genetic instruments, including weighted-median, weighted-mode, simple mode, MR-Egger, and MR-inverse variance weighted to assess consistency of our results.

As a complementary analysis, we examined PAVs, specifically missense and stop-gain or stop-loss variants with a minor allele frequency of at least 1%, in the four proteins anchored by MR evidence on cardiovascular risk factors (ANGPTL3, INHBC, PCOLCE, and PLXNB2). PAVs were identified using the Ensembl REST API.^35^ Each PAV was queried for association with the same cardiovascular risk factors (SBP, DBP, LDL-C, and TC) and clinical endpoints (CAD,^31^ T2D,^32^, stroke,^33^ and heart failure^34^) using the same GWAS summary data as in the primary analyses. PAVs were first filtered for association with at least one cardiovascular risk factor at a Bonferroni-corrected threshold accounting for the total number of PAVs tested across all four genes (p < 0.05/36 = 1.4×10 ³). PAVs passing this threshold were then examined for association with clinical endpoints, applying Bonferroni correction for the number of PAVs carried forward multiplied by the number of outcomes tested (p < 0.05/12 = 4.2 × 10 ³).

Lastly, we used peptide-level data available in 200 participants in the Cardiovascular Health Study. The Cardiovascular Health Study is a multi-center, population-based prospective cohort study designed to investigate risk factors of cardiovascular disease in older adults (≥65 years at baseline).^36,37^ Participants (n=5,201) were enrolled from 4 communities in the US (Forsyth County, North Carolina; Washington County, Maryland; Sacramento County, California; Pittsburgh, Pennsylvania) in 1989 to 1990. Additional participants (n=687) who were predominantly black were enrolled in 1992 to 1993 to increase the diversity of the cohort.

Untargeted proteomic profiling was conducted using the Proteograph XT Assay in 200 stored plasma specimens collected in 1992-1993. The Proteograph XT Assay quantifies more than 18,000 unique peptides from 5,000 proteins using five distinct nanoparticles that are enriched for different proteins.^38^ Details on SEER Proteograph have been published.^39^ For INHBC and PLXNB2, we used linear regression to test associations between the SNPs in LD with the respective missense variants (rs11172134 with rs2229357; rs28573806 with rs28379706) and both individual peptide levels and aggregate protein-level abundance.

## Data Availability

All data produced are available online. Serum proteins altered by the DASH diet have been published (Kim et al., J Am Heart Assoc. 2023), and we used data from published GWAS (please see relevant citations).

## Notes

**Funding:** HK is supported by a grant from the National Heart, Lung, and Blood Institute (NHLBI, K01 HL168232). Proteomic profiling was funded by the NHLBI (R01 HL153178). JFS was supported by NHLBI grants R01HL142599, R01HL149706, R01HL172495, R01HL172803, R01HL18197, and U24HL169645. The Cardiovascular Health Study was supported by NHLBI contracts HHSN268201200036C, HHSN268200800007C, HHSN268201800001C, N01HC55222, N01HC85079, N01HC85080, N01HC85081, N01HC85082, N01HC85083, N01HC85086, 75N92021D00006; and NHLBI grants U01HL080295, R01HL087652, R01HL103612, R01HL105756, R01HL120393, U01HL130114, and R01HL172803 with additional contribution from the National Institute of Neurological Disorders and Stroke (NINDS). Additional support was provided through R01AG023629 from the National Institute on Aging (NIA). Proteomic profiling (SEER) in CHS was supported by National Institute of Neurological Disorders and Stroke award R01NS138297.A full list of principal CHS investigators and institutions can be found at CHS-NHLBI.org. This manuscript is the result of funding in whole or in part by the National Institutes of Health (NIH). It is subject to the NIH Public Access Policy. Through acceptance of this federal funding, NIH has been given a right to make this manuscript publicly available in PubMed Central upon the Official Date of Publication, as defined by the NIH. The content is solely the responsibility of the authors and does not necessarily represent the official views of the NIH.

### Competing Interest Statement

The authors have declared no competing interest.

